# Mapping ODI onto EQ-5D-5L in Chinese Low Back Pain Patients

**DOI:** 10.1101/2024.02.20.24303104

**Authors:** Jia Li, Shuzhang Du, Chengqun Chen, Ziping Ye

**Affiliations:** Department of Pharmacy, The First Affiliated Hospital of Zhengzhou University, Zhengzhou, Henan, China; Department of Pharmacy, The First Affiliated Hospital of Zhengzhou University, Zhengzhou, Henan; School of Public Administration, Hainan University, Hainan, China

**Keywords:** Mapping, low back pain, health utilities, the 5-level EuroQol-5 Dimension, Oswestry Disability Index

## Abstract

Mapping can translate utility values from other health-related quality-of-life scales, giving researchers and policymakers more comprehensive information. The primary objective of the study is to develop mapping algorithms that convert scores from the Oswestry Disability Index (ODI) to the 5-level EuroQol-5 Dimension (EQ-5D-5L). Data for this analysis was sourced from 272 patients suffering from low back pain. The development of the mapping algorithms involved the application of three distinct regression methods across four different settings: ordinary least squares regression, beta regression, and multivariate ordered probit regression. To evaluate the internal validity of these algorithms, we adopted a ‘hold-out’ approach for predictive performance assessment. Furthermore, to discern the most effective model, three goodness-of-fit tests were employed: the mean absolute error (MAE), the root-mean-square error (RMSE), and the Spearman rank correlation coefficients between the predicted and observed utilities. The study successfully developed several models capable of accurately predicting health utilities in the specified context. The best performing models for ODI to EQ-5D-5L mapping were beta regressions. Mapping algorithms developed in this study enable the estimation of utility values from the ODI. The algorithms formulated in this study facilitate the estimation of utility values based on the ODI, providing a valuable empirical foundation for estimating health utilities in scenarios where EQ-5D data is unavailable.

## Introduction

Low back pain (LBP) is characterized by pain, muscle tension or stiffness situated beneath the costal margin and above the inferior gluteal folds, with or without sciatica[27]. LBP is a is a symptom of high prevalence and complexity, with almost everyone experiencing a brief, acute LBP during their lifetime. In 2017, the age-standardized global point incidence rate of LBP was 7.5%[28]. The incidence can be particularly high in certain professions; for instance, up to 76.0% of nurses in intensive care units may experience LBP.[24]. In China, the prevalence of LBP among workers ranges from approximately 42.7% to 72.0%, making it the leading cause of physical disability[13]. LBP is the main cause of disability life years (YLDs)[5] and is associated with a marked decrease in health-related quality of life (HRQoL) [1]. It has been observed that patients with LBP often report a lower quality of life, comparable to that experienced by individuals with life-threatening conditions[8].

Over the past two decades, China has seen a notable increase in economic evaluations with significantly more cost-per-quality-adjusted life-year (QALY) studies than cost-per-disability-adjusted life-year (DALY) studies[3]. QALY, which is largely used as clinical effectiveness indicator, is a generic preference-based measure that provides a general estimate of health outcomes and can capture survival data[25]. Globally, the EuroQol 5-dimension (EQ-5D) stands out as the most commonly used preference-based tool, and its use in clinical studies in China is gaining momentum. Despite this trend, there is a scarcity of economic evaluations specifically addressing low back pain (LBP) in the literature. This gap may be partly attributed to the limited availability of health utility data related to LBP.

Mapping or cross-walking can generate utility values from other health-related quality-of-life scales, giving researchers and policymakers a more comprehensive information[20]. The Oswestry Disability Index (ODI) is one of the most widely utilized disease-specific outcome reporting tools for measuring dysfunction associated with LBP[19; 26], which is recommended by the Chinese guideline for clinical diagnosis and treatment of non-specific LBP[10]. Given the prevalent use of disease-specific tools in clinical settings, there is a valuable opportunity to associate these condition-specific measures with utility values. Mapping algorithms, which often incorporate demographic factors such as age and gender, are a promising approach to enhance the predictive accuracy of these models. Previous studies[9; 31] confirmed that EQ-5D-5L was more consistent and sensitive in aligning with the ODI than SF-6D and EQ-5D-3L, particularly for evaluating the health status of LBP patients. Therefore, the main purpose of this study is to develop a mapping algorithm that converts HRQoL data from the ODI into EQ-5D-5L utility values. These algorithms will be instrumental in deriving in subsequent economic evaluation.

## Methods

### Data

This study used data from a cross-sectional survey[31] that focused on individuals suffering from LBP. Participants were selected based on specific inclusion criteria: they were required to be 18 years of age or older, could have lower limb pain, were only using routine painkillers for pain management, and had the ability to understand and communicate in Mandarin. Several exclusion criteria were applied. Patients were not eligible if they had concurrent infections, cancer, serious spinal cord disorders, inflammatory joint diseases, recent myocardial infarction, cerebrovascular incidents, chronic lung diseases, kidney diseases, severe mental health conditions, or if they were pregnant. The diagnosis of LBP in these patients was determined through a combination of imaging studies, physical examinations, and patient-reported symptoms related to LBP.

### Instruments

#### Oswestry Disability Index

The ODI is a widely recognized tool for assessing functional impairment related to back pain, relying on patient-reported outcomes. This questionnaire asks patients to evaluate how pain affects their ability to carry out activities across ten domains: : pain intensity, personal care, lifting, walking, sitting, standing, sleeping, sex life, social life and travelling. A higher ODI score signifies a more severe disability level associated with LBP. In our study, we employed the Chinese version of the ODI, officially provided by the Mapi Research Trust. However, as the “sex life” question is not culturally appropriate for Chinese citizens[32], the Chinese adaptation of the ODI questionnaire comprises only 9 of the original 10 items.

#### EQ-5D-5L

The EQ-5D-5L is a widely used instrument for assessing the health status of individuals on the day of interview. It encompasses five dimensions: mobility, self-care, pain/discomfort, usual activities, and anxiety/depression. Each dimension is subdivided into five levels of severity, ranging from no problem to extreme problem. The EQ-5D-5L score ranges from - 0.39 to 1, with a score of 1 represents the best possible health state[17].

### Exploratory Data Analysis

We anticipate a substantial degree of overlap between EQ-5D-5L and ODI. Table 1 illustrates the extent of this overlap, detailing the common coverage areas of both scales. While the EQ-5D-5L does not explicitly include an item on “travel,” its aspects—such as mobility, usual activities, pain/discomfort, and anxiety/depression—collectively encompass this domain. Our analysis progresses by examining the data to identify parallels between these two instruments, employing Spearman correlation coefficients to explore these similarities.

### Statistical methods

In the last ten years, a variety of regression methods have been employed in published literature for the development of mapping algorithms[20]. These algorithms aim to not only replicate mean predicted health utilities but also the distribution of the index. The suitability of these methods depends on the specific health measure being used, its distribution, and the standard tests for assessment. There are generally two primary methods for algorithm development: response mapping and mapping directly into health utility values. Response mapping analyzes health state descriptions directly with modelling techniques like multinomial logit and ordered logistic. Many researchers favor the multivariate ordered probit (MOP) method for its ability to address correlations between different dimensions[7; 21]. The more prevalent method involves mapping directly into health utility values, with ordinary least squares (OLS) regression being the most frequently used[20]. However, OLS has faced criticism for potentially producing values outside a plausible range. T To address this issue, some researchers propose using regression estimators based on the characteristics of the beta distribution, which assume that the dependent variable is confined to a range between 0 and 1 [2; 14].

#### Modelling Approaches

This research utilized three regression techniques—OLS, beta regression, and MOP—to devise mapping algorithms. The statistical analysis was executed in the R software environment, utilizing the ‘betareg’ package for the beta regression method and the ‘mvord’ package for the multivariate approach. In the OLS method, predicted utilities were constrained to a maximum of 1 for values exceeding 1. To address the constraints of the beta regression model, which cannot handle values of 0 or 1, adjustments were made for values at the lower and upper bounds of the distribution. This was achieved by rescaling the values using the following equation:

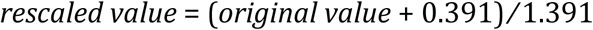

#### Choice of predictors

There are multiple approaches to incorporating the ODI into a mapping algorithm. The simplest method involves utilizing the total ODI score as a predictor for the EQ-5D-5L. Alternatively, one could opt to include the individual responses for each ODI item. Therefore, we suggested four different ways to use the ODI as explanatory variables: Model 1 – age, gender, and ODI total scores; Model 2 – age, gender, and ODI item scores; Model 3 - ODI total scores only; Model 4 - ODI item scores only. We used backward elimination to select the final variables for the regressions.

#### Goodness of fit

In this investigation, we assessed the goodness-of-fit of the fitted models by employing two widely used indicators: mean absolute error (MAE) and mean square error (MSE). Furthermore, we analyzed the Spearman rank correlation coefficients to gauge the strength of correlations between the predicted and observed values. Given the absence of external data, we used the hold-out method [4] to validate the alternative models. To carry out this procedure, we randomly partitioned all samples into two groups: 75% of the data (the estimation dataset, with N=204 patients) was utilized to construct the mapping models, while the remaining 25% (the validation dataset, with N=68 patients) was set aside for assessing the goodness-of-fit of the models. Our research adheres to the MAPS Reporting Statement guidelines [22].

## Results

### Descriptive statistics

In this survey, 300 patients with LBP were interviewed. Nevertheless, 28 patients were excluded from the analysis due to incomplete questionnaire responses (n=17) or being either too young or too old for the study (n=11). Consequently, 272 patients (91%) were incorporated into the analysis, with 204 patients (75%) randomly allocated to the estimation set and 68 patients (25%) to the validation set. Table 2 illustrates the demographic and clinical attributes of patients within the estimation and validation sets.

Table 3 provides the summary statistics of the EQ-5D-5L and ODI in both the estimation and validation sets, along with a more detailed breakdown of the ODI dimensions. In the estimation and validation set, the mean (SD) EQ-5D-5L index was 0.60 (0.34) and 0.61 (0.32), and the mean (SD) of ODI total score is 0.33 (0.21) and 0.34 (0.21), respectively, which indicates little difference in these two data sets.

### Overlap between ODI and EQ-5D-5L

The overlap between ODI and EQ-5D-5L is shown in Table 4. Among the nine dimensions of ODI, personal care, lifting, walking, social life and traveling have stronger correlation with EQ-5D-5L subscale (r≥0.5, highlighted in bold). At the same time, utility score shows strong correlation with seven dimensions in ODI except sitting and sleeping (r≥0.5). The correlations all reached statistical significance (p < 0.01).

### Model estimates

Table 5 displays the parameter estimates for each of the mapping algorithms. It is critical to acknowledge that, for OLS models, predicted values exceeding 1 were capped at 1. When dealing with predictions derived from beta regression models, it’s essential to revert them to the original scale, which spans from -0.391 to 1. This can be accomplished using the following equation: *original value = (predictions * 1* .39*1) - 0.391*

According to the primary measure of prediction accuracy, which is MSE, beta regressions provided the best predictions (MSE: 0.0176-0.0274) for all four settings. Nevertheless, in regards with the spearman rank correlation coefficients, the OLS models (OLS1, OLS3) performed better than the corresponding beta regressions. However, MOP models resulted in relatively poor predictions with an MSE ranging from 0.0324 to 0.0362. The results obtained from the MAE were comparable with the MSE.

### Optimal Mapping Functions

Mapping equations and validation results to the optimal methods of 4 different settings are reported in Table 6 and Table 7. None of the models found gender to be a significant variable, and the dimensions of lifting, sitting, standing, and sleeping were not included in any of the regressions. The validation regressions demonstrate similar results with the estimation model, indicating the robustness of the models. Scatter plots in Figure 1 show the similarity between the distributions of observed and predicted utilities in the selected models.

**Fig. 1.**
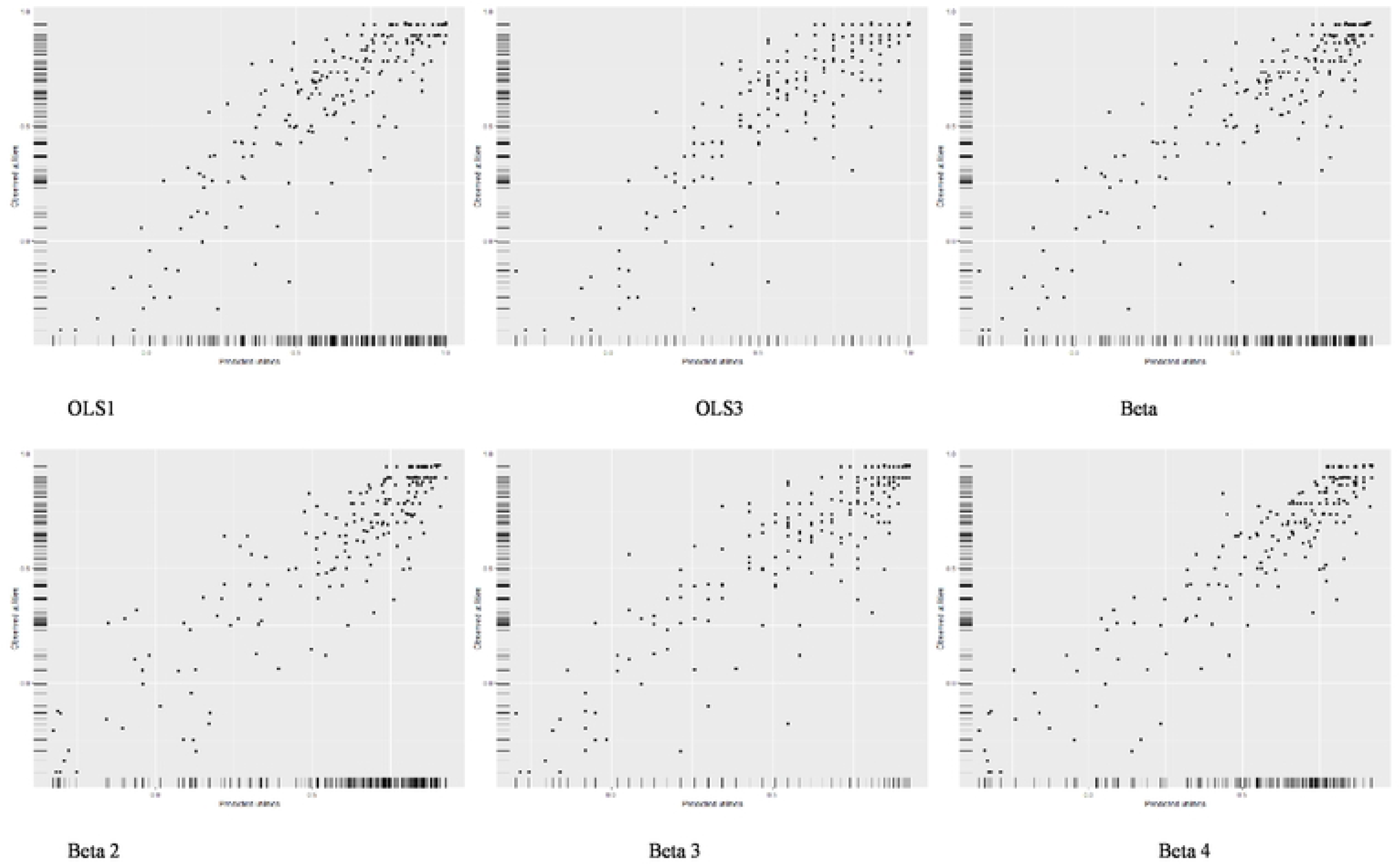
The scatter plots between observed and predicted utilities of selected models.

## Discussion

In this study, mapping algorithms were developed using a cross-sectional dataset from a tertiary hospital in China. Three established statistical methods, OLS, beta regression, and MOP regression, were used to create the mapping algorithms. This article has the potential to facilitate the evaluation of cost-effectiveness analyses (CEA) and broader impacts of treatments for individuals suffering from LBP. We proved that beta regressions outperformed the commonly used econometric mapping approaches (OLS and MOP methods).

Comparing the performance of this study with previous mapping literature on LBP patients is challenging due to the use of different disease-related or preference-based instruments. Moreover, few studies have been published on developing mapping algorithms for health utilities in LBP patients[15; 18; 23]. Khan and colleagues attempted to estimate mapping algorithms from the Roland Morris Questionnaire, another disease-specific patient reporting outcome tool for measuring back pain-related dysfunction, to EQ-5D and SF-6D[18]. They showed that the mapping algorithms from the Roland Morris Questionnaire to EQ-5D had a range of MSE of selected models between 0.0380 to 0.0397, which were larger than the goodness-of-fit outcomes reported in this study. The mapping algorithms derived from the Roland Morris Questionnaire showed significantly smaller MSE when mapping to SF-6D compared to EQ-5D, likely due to the stronger correlation between the Roland Morris Questionnaire and SF-6D. In this study, we used the ODI, which was found to have a stronger correlation with EQ-5D-5L than SF-6D, justifying the target measurements used in this research.

To assess the predictive accuracy of the mapping algorithms formulated in this study, we estimated the MSE, MAE, and Spearman rank correlation coefficient. However, the MOP models showed higher prediction errors in comparison to beta regression and OLS, which is contrary to the findings of Patton and colleagues[21]. Several reasons may explain this disagreement. Firstly, the optimal method for mapping algorithms may differ for different diseases. Additionally, the dataset’s sample size in their study, which was used to develop the mapping algorithms, was small, potentially influencing the statistical power. Beta distributions have been proposed as a flexible option for modeling health utilities [2], a finding that aligns with our results. Although we cannot draw general conclusions regarding recently proposed methods for estimating mapping algorithms, our study underscores the potential advantages of such approach and emphasize the importance of considering a wide range of candidate regression models during mapping construction.

There are growing numbers of studies estimating mapping functions from disease-specific measures to EQ-5D. For instance, He and colleagues proved that the OLS performed better than GLM, CLAD, and Tobit model regression in mapping the cancer-specific instrument (FACT-G) to a preference-based measure (EQ-5D-3L)[11]. Xu et al. formulated algorithms to map the EORTC QLQ-C30 (QLQ-C30) onto EQ-5D-5L within a patient sample diagnosed with lymphomas[30]. More mapping algorithms were developed including psoriasis disability index, Haem-A-QoL scores, Parkinson’s Disease Questionnaire[6; 12; 29].

To accurately interpret the results, some limitations should be taken into consideration. Firstly, the study had a sample size of 300 patients, and the analyses were only performed on the 272 patients who completed all the questionnaires. As this was a convenience sample, the results may not be representative of all patients with LBP. Secondly, the validation results may not reflect the true performance of the model on unseen data, as an external dataset for validation was not available. Therefore, it is strongly recommended to use an external dataset for validation. Finally, it is important to note that mapping algorithms should be regarded as a secondary option for generating health utilities from non-preference-based, disease-specific quality of life instruments.

## Conclusion

Recently, mapping algorithms that link disease-specific instruments to generic measures of HRQL have played a vital role in the development of cost-effectiveness studies. This study, to the best of the authors’ knowledge, is the first to have developed a mapping algorithm between the ODI and EQ-5D-5L. The data used to develop the algorithms were collected from a cross-sectional study of LBP patients in a tertiary referral hospital in China. Several algorithms were developed, and it was found that the beta regression method had better predictive performance. Although the estimation data had a small sample size, the developed algorithms demonstrated comparable predictive performance to existing mapping studies. The availability of these algorithms may aid their use in applied CEA studies.

## Data Availability

All relevant data are within the manuscript and its Supporting Information files. The raw data that used in this study has been published on https://hqlo.biomedcentral.com/articles/10.1186/s12955-019-1137-6.

## Acknowledgments

This work was supported by the National Key R&D Program of China “Active Health and Aging Response” (2020YFC2005500)

## Ethics approval

Low back pain patients were recruited at the General Hospital of Shenyang Military Area Command. Informed consent was obtained from each patient. The Ethics Committee of the General Hospital of Shenyang Military Area Command granted ethics approval (Code of Ethics: K (2017)22).

## Consent for publication

Written consent was acquired from each patient.

## Data availability statement

The datasets generated during and/or analyzed during the current study are available from the corresponding author on reasonable request.

---

**Table 1** Overlap between the descriptive systems of the EQ-5D-5L and the ODI.

**Table 2** Patient characteristics.

**Table 3** Descriptive statistics for health utility and HRQoL variables.

**Table 4** Spearman rank correlations of ODI dimension scores with EQ-5D-5L values.

**Table 5** Model estimates for mapping from ODI to EQ-5D-5L.

**Table 6** Mapping equations for the optimal methods.

**Table 7** Model validations for the optimal methods.

## Notes

### Competing Interest Statement

The authors have declared no competing interest.

### Funding Statement

Yes

### Author Declarations

Low back pain patients were recruited at the General Hospital of Shenyang Military Area Command. The Ethics Committee of the General Hospital of Shenyang Military Area Command granted ethics approval (Code of Ethics: K (2017)22).

